# The Lifelines COVID-19 Cohort: a questionnaire-based study to investigate COVID-19 infection and its health and societal impacts in a Dutch population-based cohort

**DOI:** 10.1101/2020.06.19.20135426

**Authors:** Kate Mc Intyre, Pauline Lanting, Patrick Deelen, Henry Wiersma, Judith M. Vonk, Anil P.S. Ori, Soesma A. Jankipersadsing, Robert Warmerdam, Irene van Blokland, Floranne Boulogne, Marjolein X.L. Dijkema, Johanna C. Herkert, Annique Claringbould, Olivier Bakker, Esteban A. Lopera Maya, Ute Bültmann, Alexandra Zhernakova, Sijmen A. Reijneveld, Elianne Zijlstra, Morris Swertz, Sandra Brouwer, R. van Ooijen, Viola Angelini, Louise Dekker, Anna Sijtsma, Sicco A. Scherjon, Jackie Dekens, Jochen O. Mierau, H. Marike Boezen, Lude Franke

## Abstract

The COVID-19 pandemic has affected billions of people around the world not only through the infection itself but also through its wider impact on public health and daily life. To assess the effects of the pandemic, a team of researchers across a wide range of disciplines developed and implemented the Lifelines COVID-19 questionnaire, leading to the development of the Lifelines COVID-19 cohort. This cohort is recruited from participants of the Lifelines prospective population cohort and the Lifelines NEXT birth cohort, and participants were asked to fill out detailed questionnaires about their physical and mental health and experiences on a weekly basis starting in late March of 2020 and on a bi-weekly basis staring in June 2020. The Lifelines region covers the three Northern provinces of the Netherlands— Drenthe, Groningen and Friesland—which together account for ∼10% of the Dutch population. To date, >70,000 people have responded to the questionnaires at least once, and the questionnaire program is still ongoing. Data collected by the questionnaires will be used to address four aspects of the outbreak: (1) how the COVID-19 pandemic developed in the three northern provinces of the Netherlands, (2) which environmental risk factors predict disease susceptibility and severity, (3) which genetic risk factors predict disease susceptibility and severity and (4) what are the psychological and societal impacts of the crisis.

**Informed consent:** All Lifelines and Lifelines NEXT participants have provided informed consent that provide the opportunity for add-on research.

**Research involving human participants:** Both the Lifelines and the Lifelines NEXT studies were approved by the ethics committee of the University Medical Center Groningen.

## Introduction

COVID-19—the disease caused by infection with the novel coronavirus SARS-CoV-2—has impacted the lives and health of billions of people around the world. Due to the absence of a vaccine, lack of effective antiviral medication and limited understanding of the virus itself, massive efforts have been undertaken by most governments to slow the growth rate of infections through public health measures that have included tracking and testing, shutting down of public life, social distancing policies and stay-at-home orders. These steps have had a huge impact on public well-being, health, the economy (including employment and working conditions) and daily life. The COVID-19 pandemic thus has the potential to exacerbate health and economic inequalities because the concentration of health and labour market disadvantages will be among vulnerable people, such as those with chronic diseases and/or mental health problems. Psychological distress about the pandemic may also have an impact on mental health, as will social isolation. For youth, the disease risks due to COVID-19 are thought to be small, but the mental health effects of the threat of the pandemic and the associated measures such as school closures may be considerable. The pandemic may also result in unemployment, which could widen socio-economic inequalities in the near future depending how social security systems are able to react and protect. The economic impact will thus further increase the uncertainty and psychological distress experienced by many people.

The effects of the COVID-19 pandemic will therefore be multiple: there will be the impact of the infection itself and the broader societal and health impacts. To identify genetic and environmental risk factors for COVID-19 and address the medical, social and psychological aspects of the pandemic, we developed and implemented a COVID-19 questionnaire, leading to the development of the Lifelines COVID-19 cohort. The questionnaire collects data about COVID-19–related symptoms, current health issues and societal impacts from participants recruited from the Lifelines population cohort [1] and the Lifelines NEXT birth cohort [2], which both monitor the health of the northern Dutch population.

Through a questionnaire that is sent out (bi-)weekly, the project gathers information about the symptoms of COVID-19, associated comorbidities and environmental factors, changes in work and employment, corona-related worries, loneliness and the mental health and societal impacts of the pandemic. In addition, all participating parents are asked about their children’s well-being and Lifelines NEXT parents receive detailed questions about COVID-19-related symptoms expressed by their children. The project has been able to ask questions on a weekly basis since March 30, 2020 that have assessed the evolution of the COVID-19 outbreak and its influence on health in the three northern provinces of the Netherlands.

The Lifelines COVID-19 cohort is a collaboration between researchers from Lifelines; Lifelines NEXT; the University Medical Center Groningen Departments of Epidemiology, Genetics, Nephrology, Psychiatry and Health Sciences; the University of Groningen Faculty of Economics and Business and Faculty of Behavioural and Social Sciences; and the Aletta Jacobs School of Public Health. The data collected by the questionnaires will be used to address four aspects of the outbreak: (1) how the COVID-19 pandemic developed in the three northern provinces of the Netherlands, (2) which environmental risk factors predict disease susceptibility and severity, (3) which genetic risk factors predict disease susceptibility and severity and (4) what are the psychological and societal impacts of the crisis.

### COVID-19 in the Netherlands and the Northern provinces

The first three official cases of COVID-19 in the Netherlands were registered on February 27, 2020 [3]. By March 24, 2020, the number of cases diagnosed per day had risen to 1,126, with 750-1400 new cases per day being registered by the Rijksinstituut voor Volksgezondheid en Milieu (RIVM) through April 24, 2020 (Figure 1B). Over this period, the number of deaths officially attributed to COVID-19 rose to ∼150 per day. The rapid rise in case numbers in early March led the Dutch government to shut down primary and secondary schools, bars and restaurants, sporting facilities and other public spaces on March 15, 2020, followed by a more extensive shut-down of public life in the weeks that followed (see Figure 1A for major events). However, the three northern Dutch provinces did not follow the national trends. COVID-19 appeared later in the northern provinces (first reported cases: Drenthe - March 1, 2020, Friesland - March 10, 2020, Groningen - March 11, 2020), ramped up more slowly and never reached the same incidence of cases or infection rates (see Reproductive values in Figure 1C). In the Netherlands, up until June 9, 2020, a total of 47,903 SARS-CoV-2 infections were reported, and the number of COVID-19 hospitalizations and deaths are 11,800 and 6,031, respectively. For the Northern provinces combined, the number of infections, hospitalizations, and deaths over the same period were 1,491 (Drenthe=523, Groningen=352, Friesland=616), 320 (Drenthe=116, Groningen=74, Friesland=130), and 122 (Drenthe=40, Groningen=17, Friesland=65), respectively (see Supplementary table 1). Figure 2 shows the geographical distribution of the number of hospitalizations across all Dutch provinces through mid-May.

**Figure 1.**
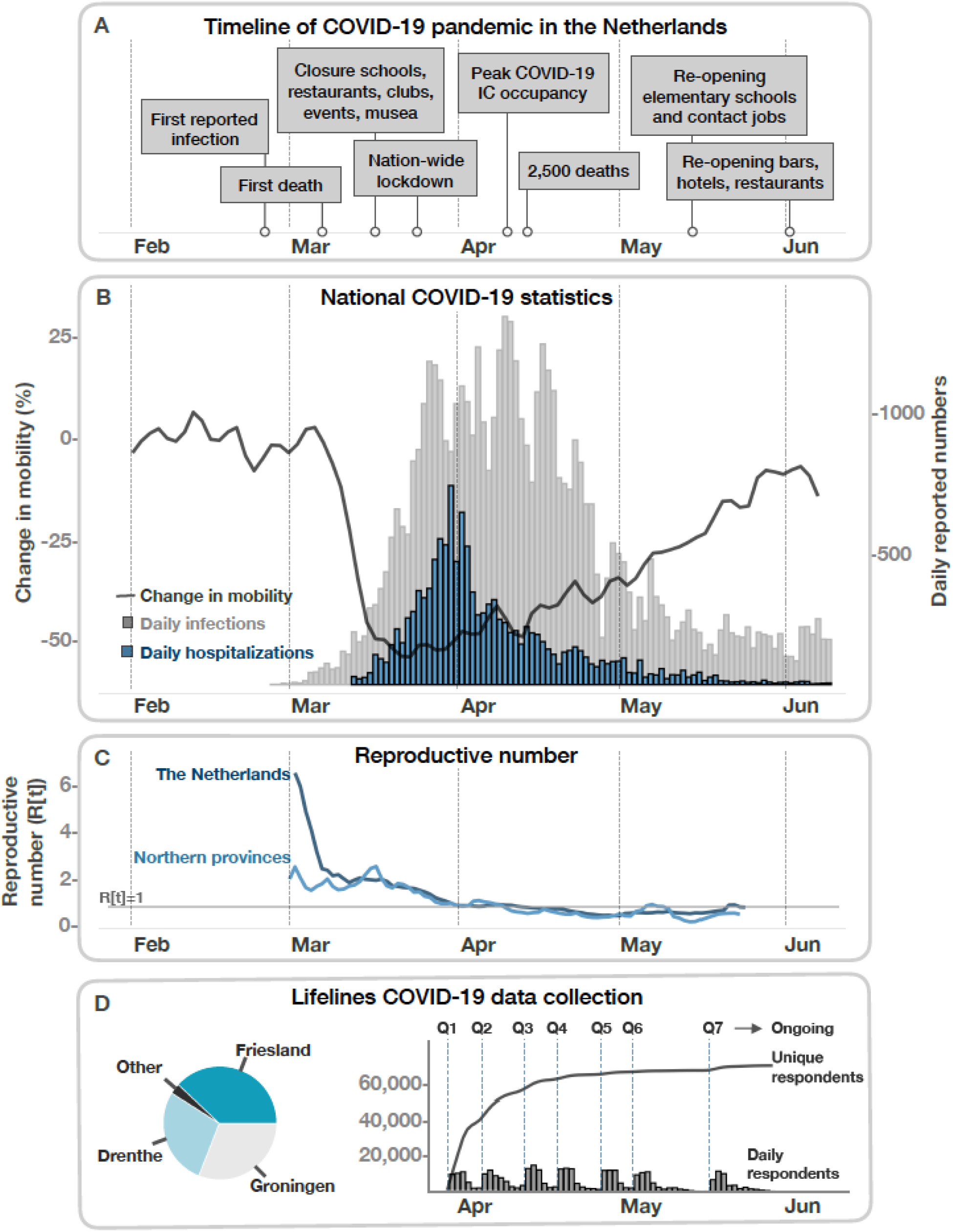
Timeline of the COVID-19 pandemic in the Netherlands and Lifelines data collection. 1A. Important events of the pandemic in the Netherlands from February to June 2020. 1B. Daily reported positive infections (grey) and hospitalizations (blue) visualized alongside the change in mobility (black) in the Netherlands. Mobility is quantified using Apple Maps Request data (https://www.apple.com/covid19/mobility) with the change over time normalized to February 1, 2020. Change in mobility indicates the percentage change in overall requested driving directions by users of Apple Maps. COVID-19 daily infections and hospitalizations are derived from the CoronaWatchNL github account (https://github.com/J535D165/CoronaWatchNL) and are based on reported numbers from the RIVM. 1C. The reproductive number in the Netherlands and the three Northern provinces over time. The R(t) is calculated based on incident cases (new positive PCR tests) including healthcare workers and cases appertaining to local outbreaks. National and regional R(t) values in the early phase of the pandemic are not directly comparable, since testing among healthcare workers was more widely adopted early on in the Northern provinces. 1D. Overview of the Lifelines COVID-19 data collections. The pie chart on the left shows the proportion of participants for each province. The first weekly COVID-19 questionnaire (Q1) was sent out on March 30, 2020. Based on Q1-7, 71,800 unique respondents have filled out at least one questionnaire. From Q7, assessments are biweekly.

**Figure 2.**
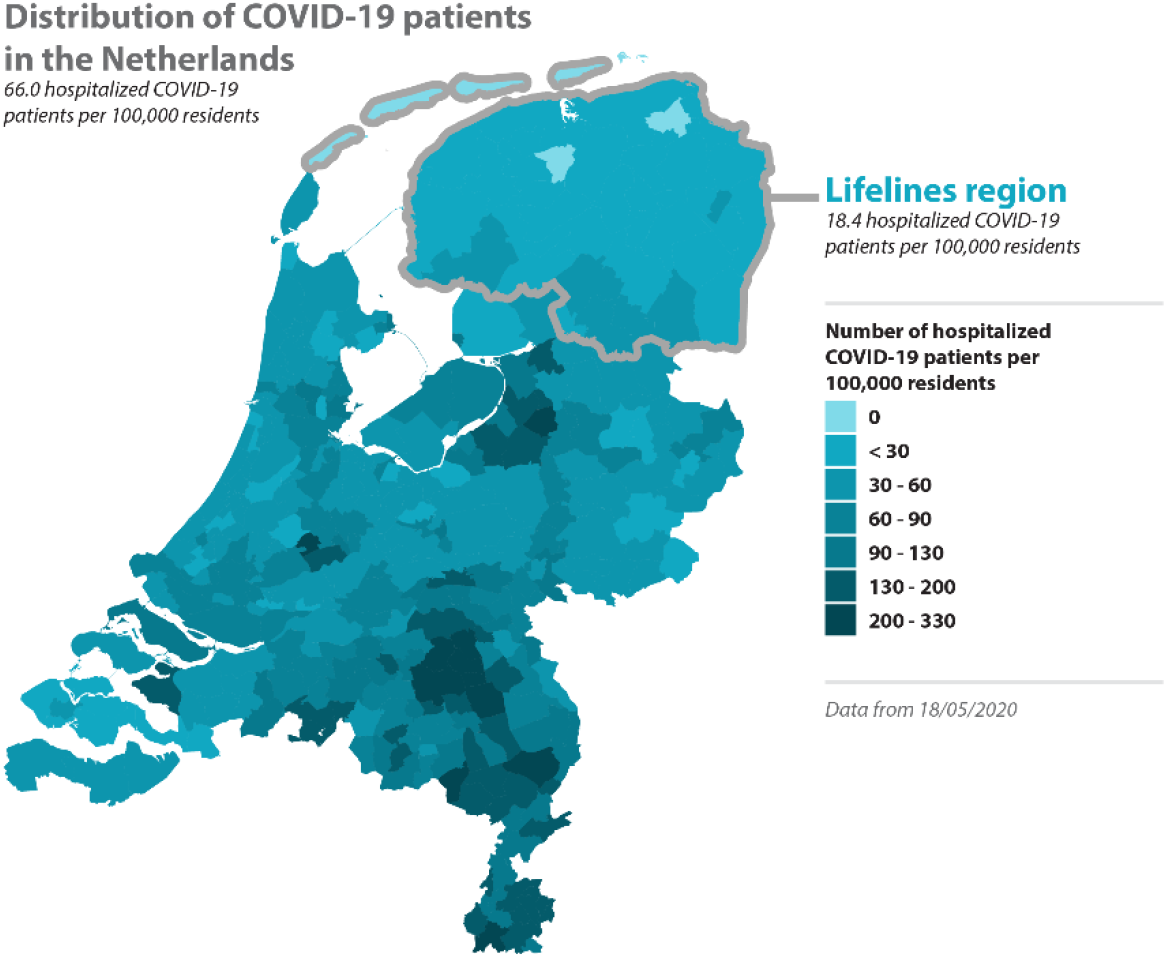
Distribution of hospitalization across Dutch municipalities. The number of hospitalizations per municipality, as reported by the RIVM, were integrated with a geographical map of the Netherlands. For each municipality, the cumulative number of COVID-19 hospitalizations per 100,000 residents is shown. The Lifelines region is outlined in grey. This data was downloaded on May 18, 2020.

**Figure 3.**
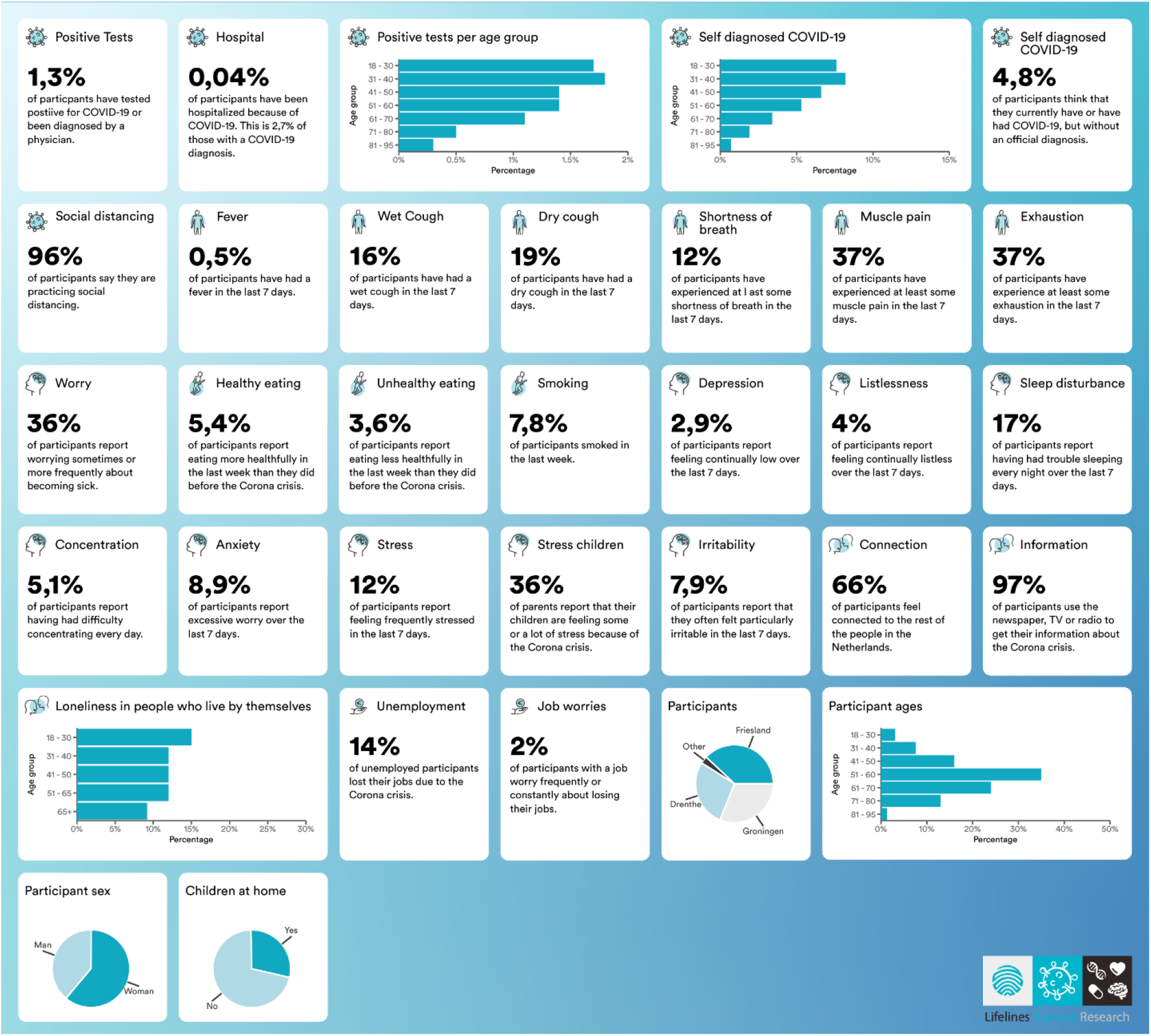
Communicating COVID-19 cohort results to the public through the Coronabarometer. Snapshot of the Coronabarometer (https://coronabarometer.nl/, *in Dutch*), which is updated after every questionnaire round to present the most recent findings of the Lifelines COVID-19 questionnaire in a format accessible by the public. The website is now interactive to enable users to look at trends over time and compare variables.

Multiple factors have been proposed to explain why the outbreak started later in the Northern provinces, and why its spread was more contained [4][5]. First, Drenthe, Groningen and Friesland together are the least-populated region of the Netherlands, accounting for ∼10% of the Dutch population, and contain the fewest urban centres. In addition, the early arrival and spread of infection in the southern Dutch province of North-Brabant seems to have originated in travel to and from Northern Italy during the school holidays from February 22-March 1, 2020, with the spread of the infection in the southern region then further facilitated by personal contact during regional carnival celebrations. In contrast, the school holidays fell earlier for the Northern provinces, from February 15-22, 2020, which suggests that northerners who travelled to Italy during the vacation had returned before the major expansion of the outbreak in Northern Italy [6]. Nor is carnival widely or generally celebrated in the Northern provinces. Other factors may also play a role, e.g. the local testing regime for COVID-19 was stricter, more tests were available in the Northern provinces and healthcare workers had their movements restricted once it became clear that cross-regional travel was a source of new infections [5].

The later arrival of COVID-19 to the North meant that the national steps taken to bring down the infection rate were in place before the outbreak had really taken hold in the region covered by Lifelines, where the caseload has so far remained the lowest in the country (Figure 2). It was against this backdrop that the Lifelines COVID-19 research was developed and implemented.

## Methods

### Participant pool

Participants of the Lifelines COVID-19 cohort are recruited from the Lifelines population cohort and the Lifelines NEXT birth cohort. Lifelines is a prospective population cohort following ∼167,000 people in the three Northern provinces of the Netherlands [1]. The cohort was established in 2006 and collects detailed information about participants via extensive questionnaires and medical examinations. As a population cohort, Lifelines is ideal for investigating common diseases and their relation to lifestyle and environmental factors in a cross-section of the population of the three northern provinces, and the cohort has been shown to be representative of the Northern Dutch population [7]. Lifelines was also designed to recruit multiple participants within families to produce a multi-generational cohort that could map individual and community health across life-course [1]. Since 2016, Lifelines NEXT has been recruiting an additional generation through inclusion of mother-baby pairs, with partners also invited to participate to generate parent-baby trios [2]. In addition, sub-cohorts within Lifelines such as Lifelines DEEP [8] and Lifelines DAG3 have collected much more detailed biological measurements, including genotype, metagenomics, metabolomics and transcriptomics. Lifelines data can also be linked to the administrative records held by Statistics Netherlands (https://www.cbs.nl/en-gb), which include health-related records on mortality, hospital admissions and healthcare costs, as well as data on employment status, income, wealth and other socio-demographic characteristics. This offers a huge potential for societally relevant research about, for instance, the long-term impacts of the SARS-CoV-2-outbreak on socioeconomic disparities in work and health.

### Recruitment strategy and questionnaire timeline

To recruit participants for the Lifelines COVID-19 cohort, Lifelines and Lifelines NEXT invited their participants digitally to fill out the questionnaires. All Lifelines participants over the age of 18 for whom an email address is known received a link to the digital COVID-19 questionnaire according to the schedule shown in Figure 1D. The digital invitations were valid for three weeks, and the date on which the questionnaire was completed is registered. All eligible Lifelines participants are invited to participate in each questionnaire round (Q1-Q7 in Figure 1D). Since Lifelines NEXT is an ongoing project in which new participants are still being included, the number of NEXT participants invited increased with each new questionnaire. Invited participants are free to choose if they want to fill in the questionnaire, and the cohort population consists of all those who have filled out at least one questionnaire over the time period of the project.

On March 30, 2020, all Lifelines and Lifelines NEXT participants were invited to participate in the first COVID-19 questionnaire round (Q1), with new invitations to participate sent out weekly following the timeline in Figure 1D. Starting in the week of March 27, 2020, an additional questionnaire about children’s health and symptoms was sent to the participants of the Lifelines NEXT (>300 participants). Questionnaires were sent out weekly through the week of May 18, 2020, and at bi-weekly intervals thereafter. As of writing, the project is set to continue into at least the summer and fall of 2020, which means that recruitment and data gathering are still on-going.

### The Lifelines COVID-19 Questionnaire Contents

The questionnaire includes modules on socio-demographic parameters, chronic diseases, COVID-19 infection, general health and symptoms, medication use, mental health/well-being of the respondent and of children and young adults in their family, corona-related well-being, social life, social relations and lifestyle (see Table 1 for questions). For participants answering a subsequent version of the questionnaire, these questions are related to their experience in the period since the previous questionnaire, either 7 or 14 days depending on the timing of questionnaires. Data collected by the questionnaires is also linked to data for participants already stored in Lifelines (e.g. height, educational background and many kinds of data from biological samples). Additional questions and question modules have been added as the questionnaire program progressed.

**Table 1.**
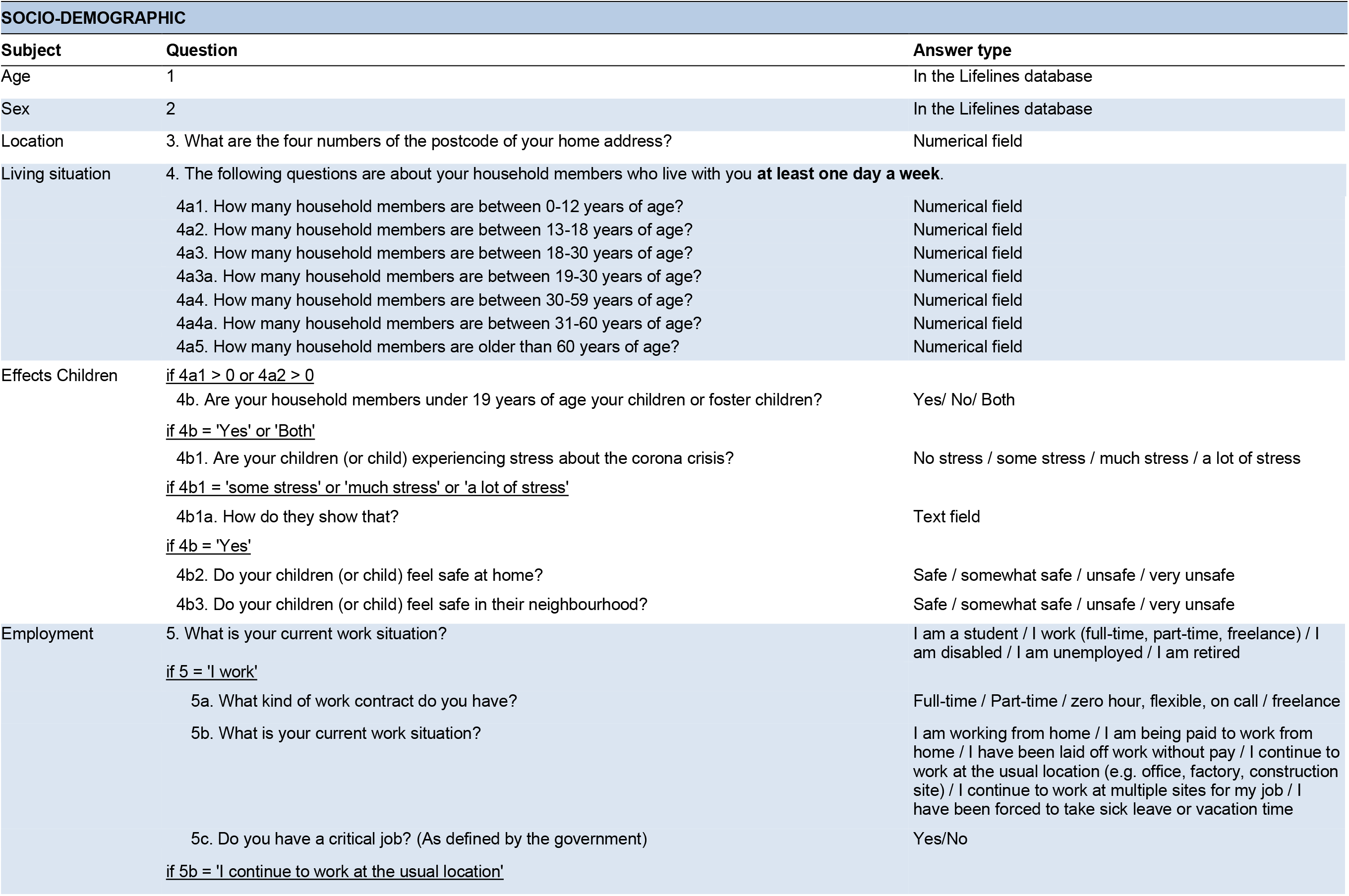
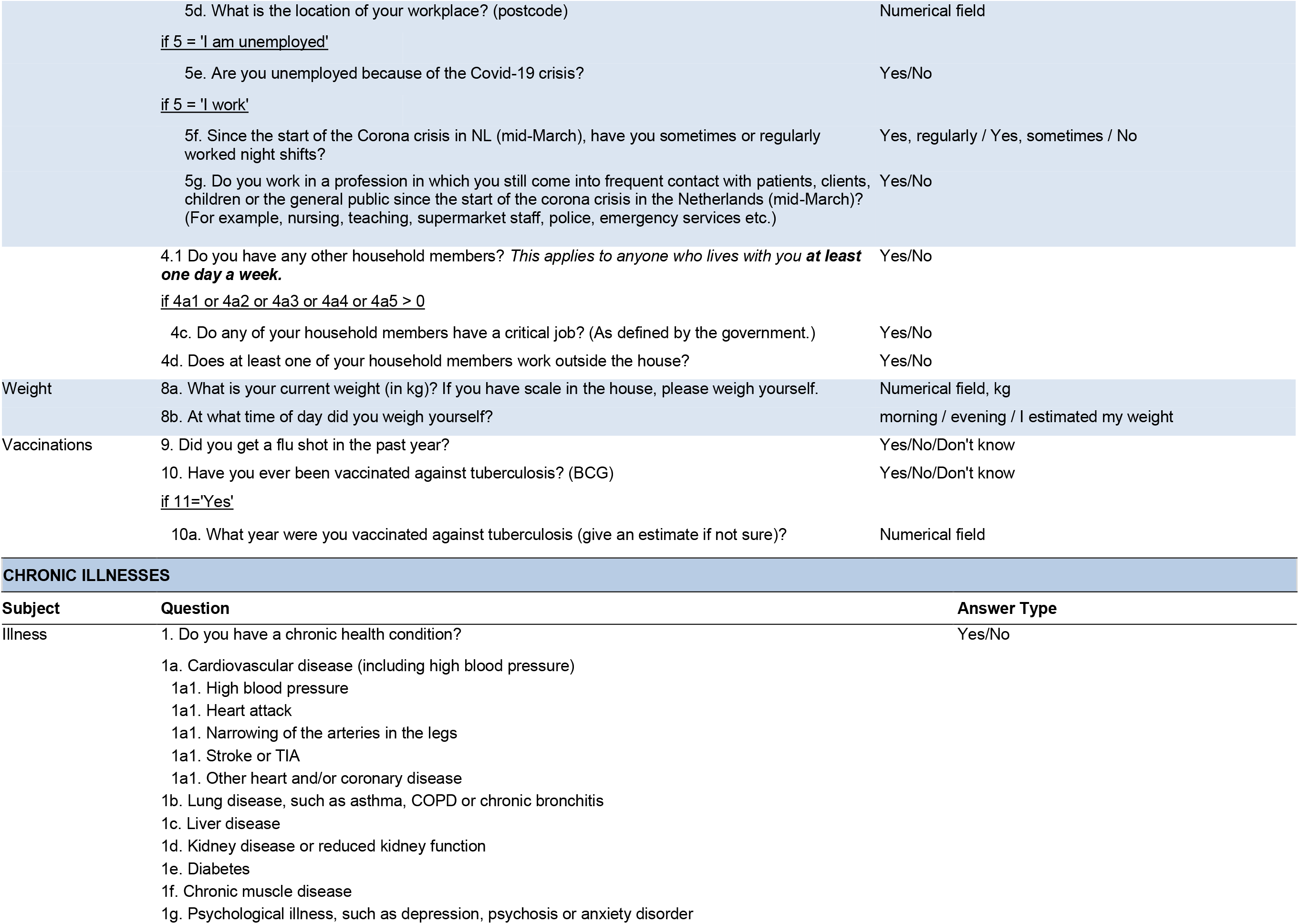
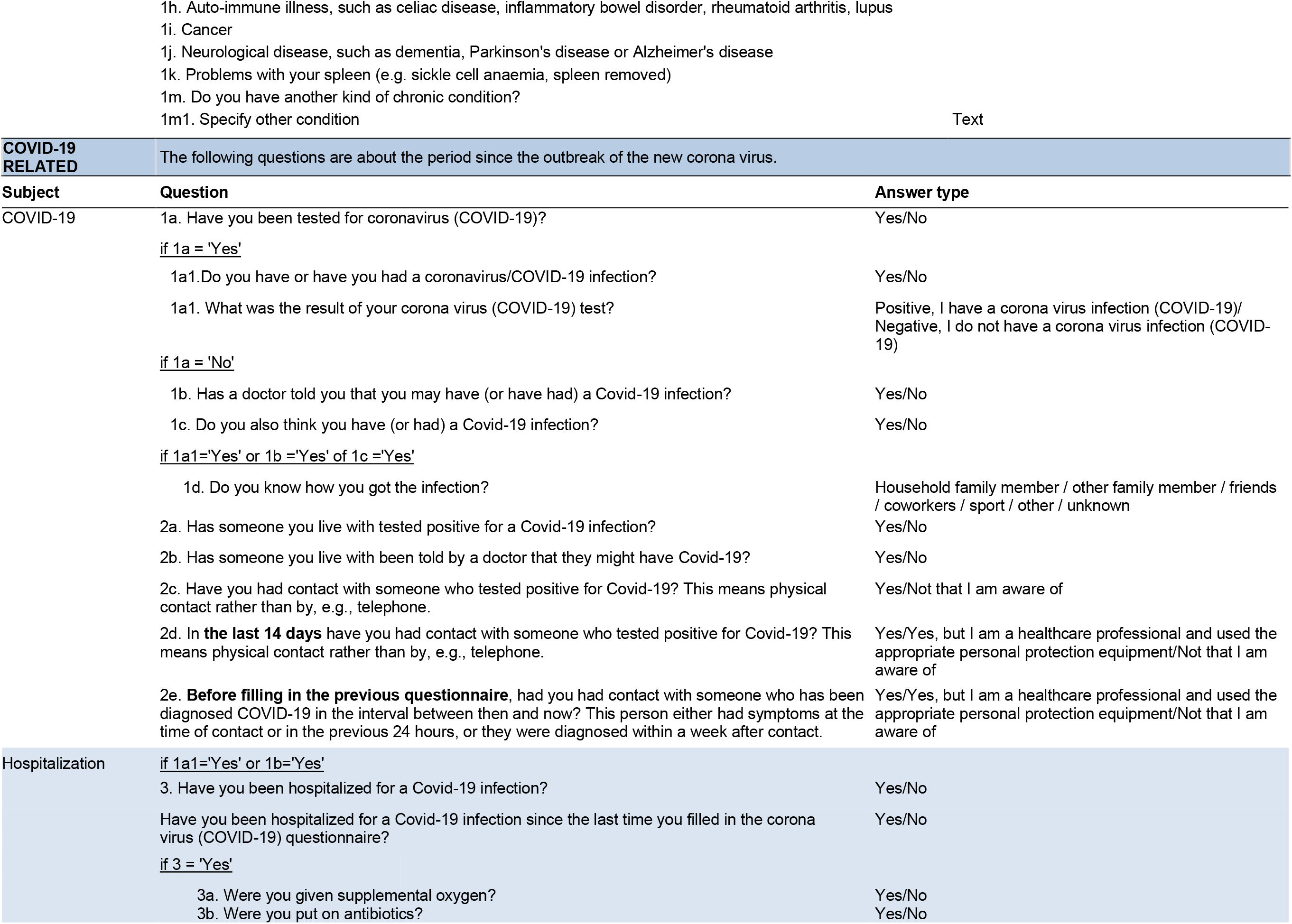
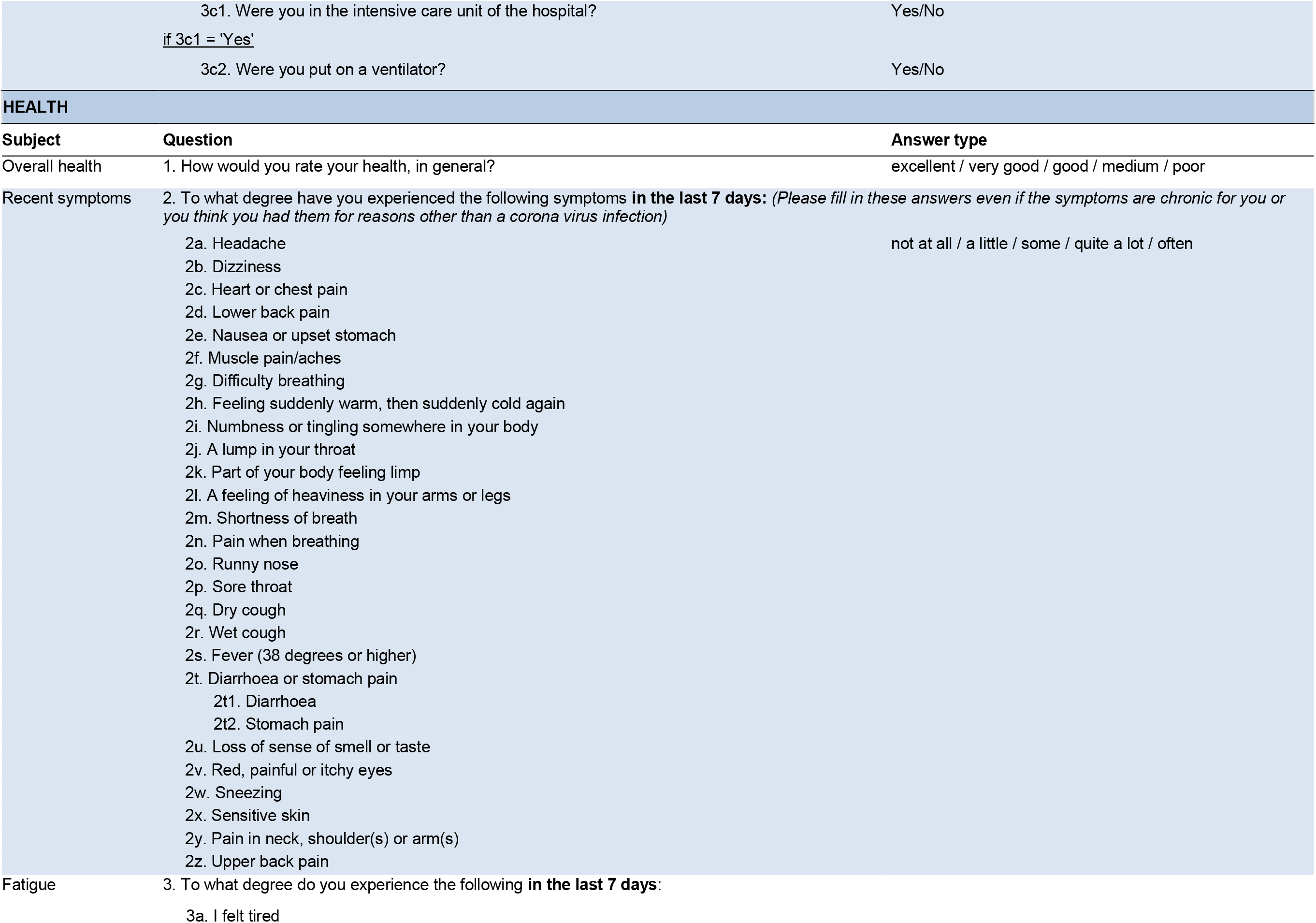
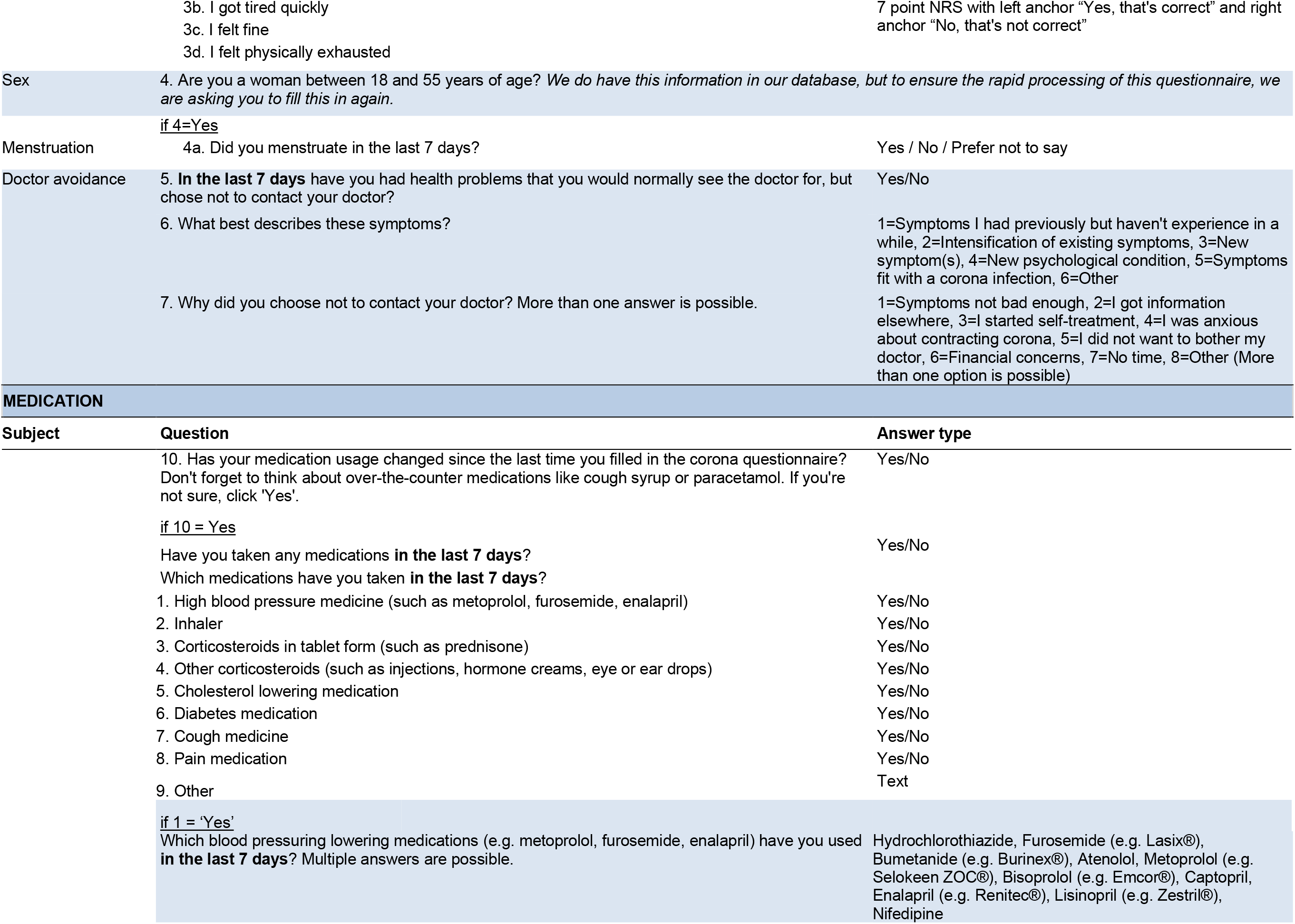
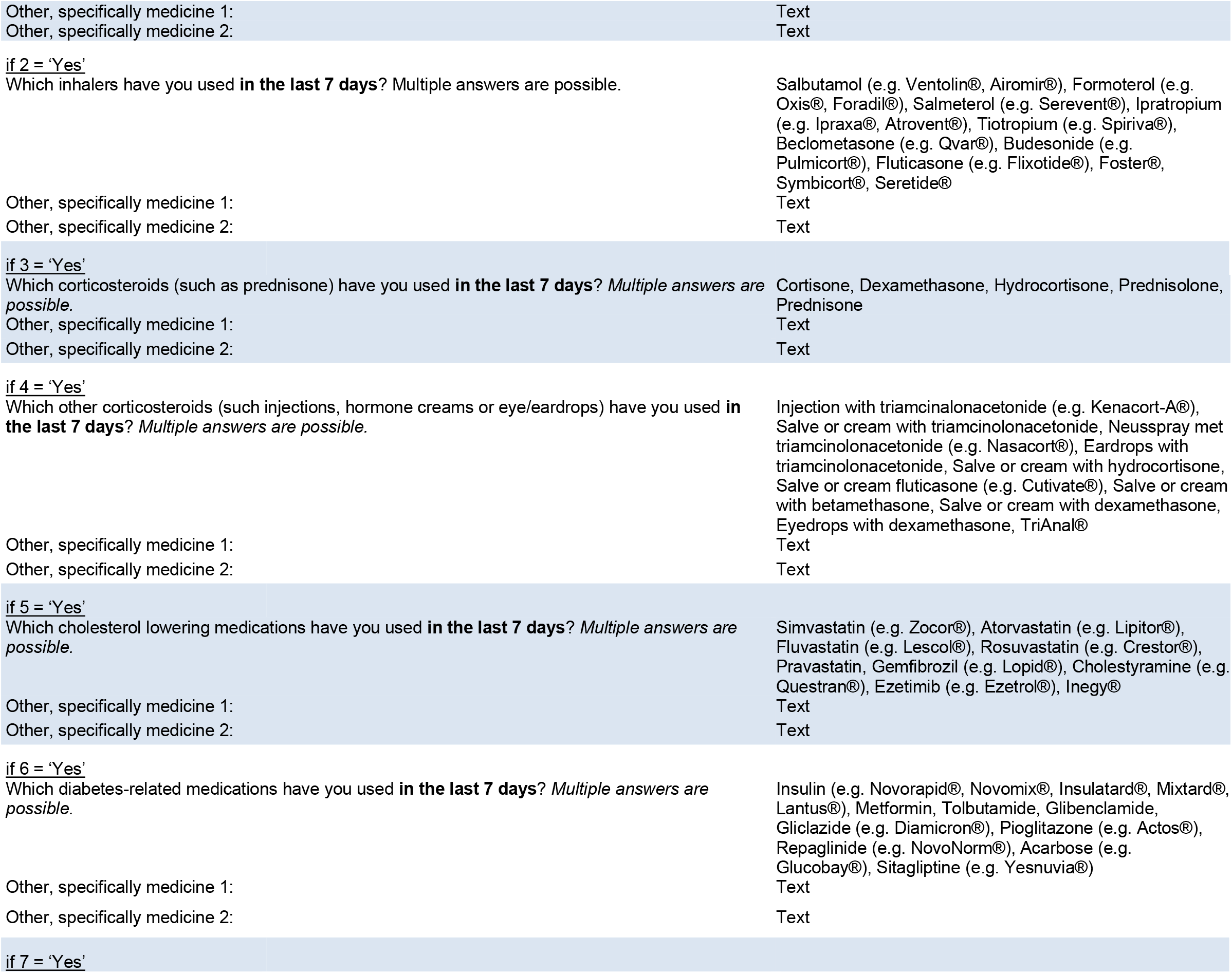
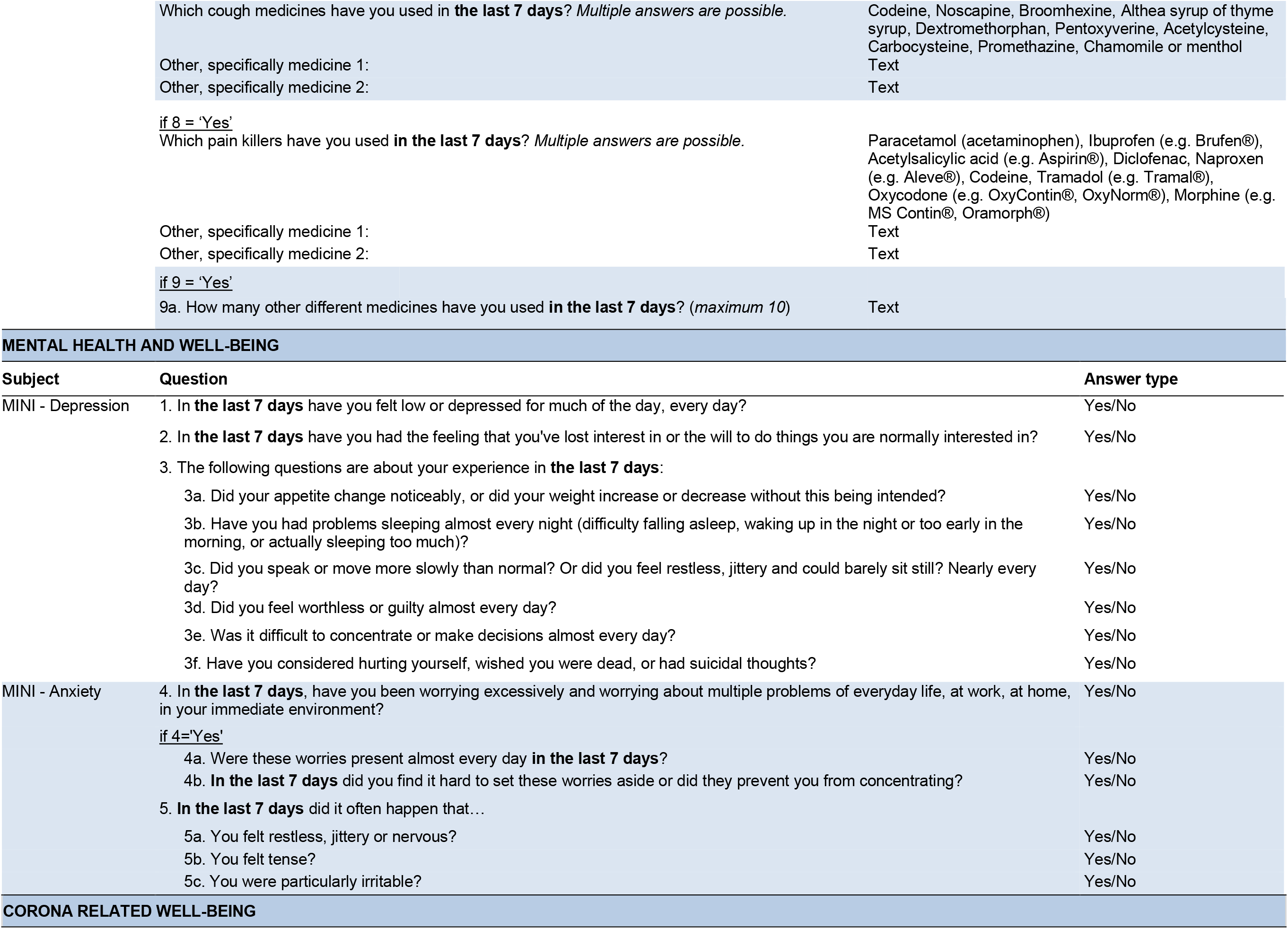
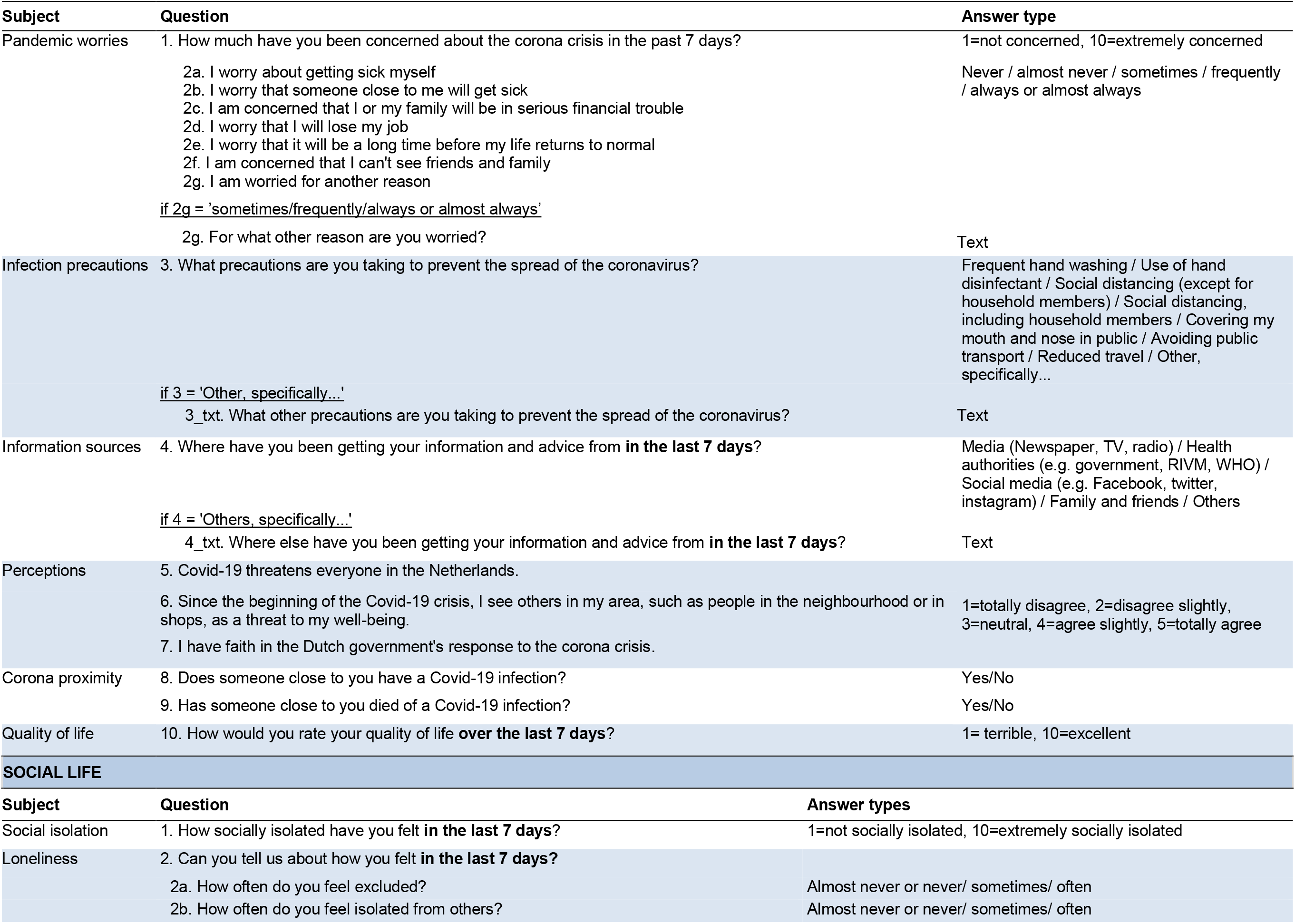
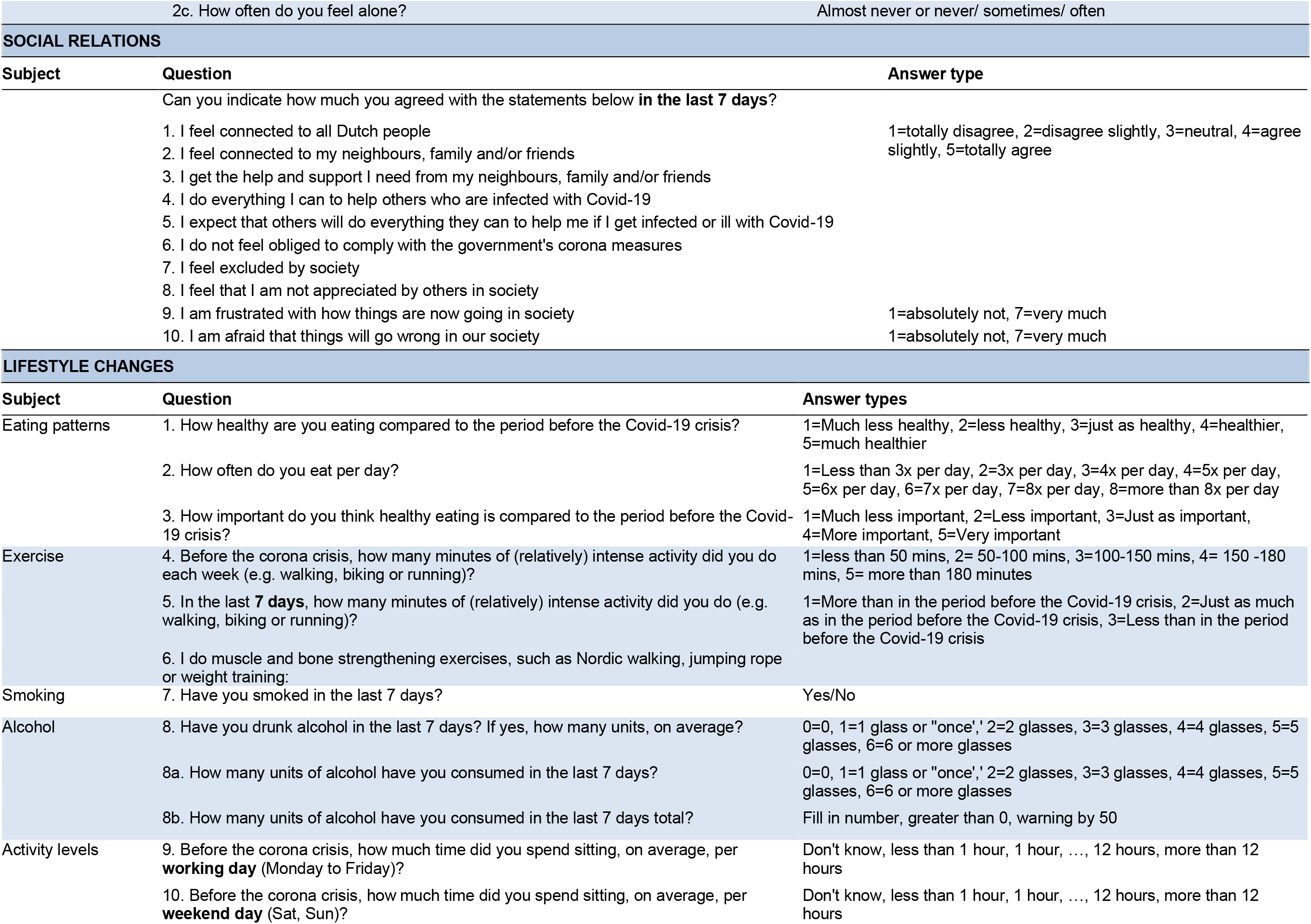
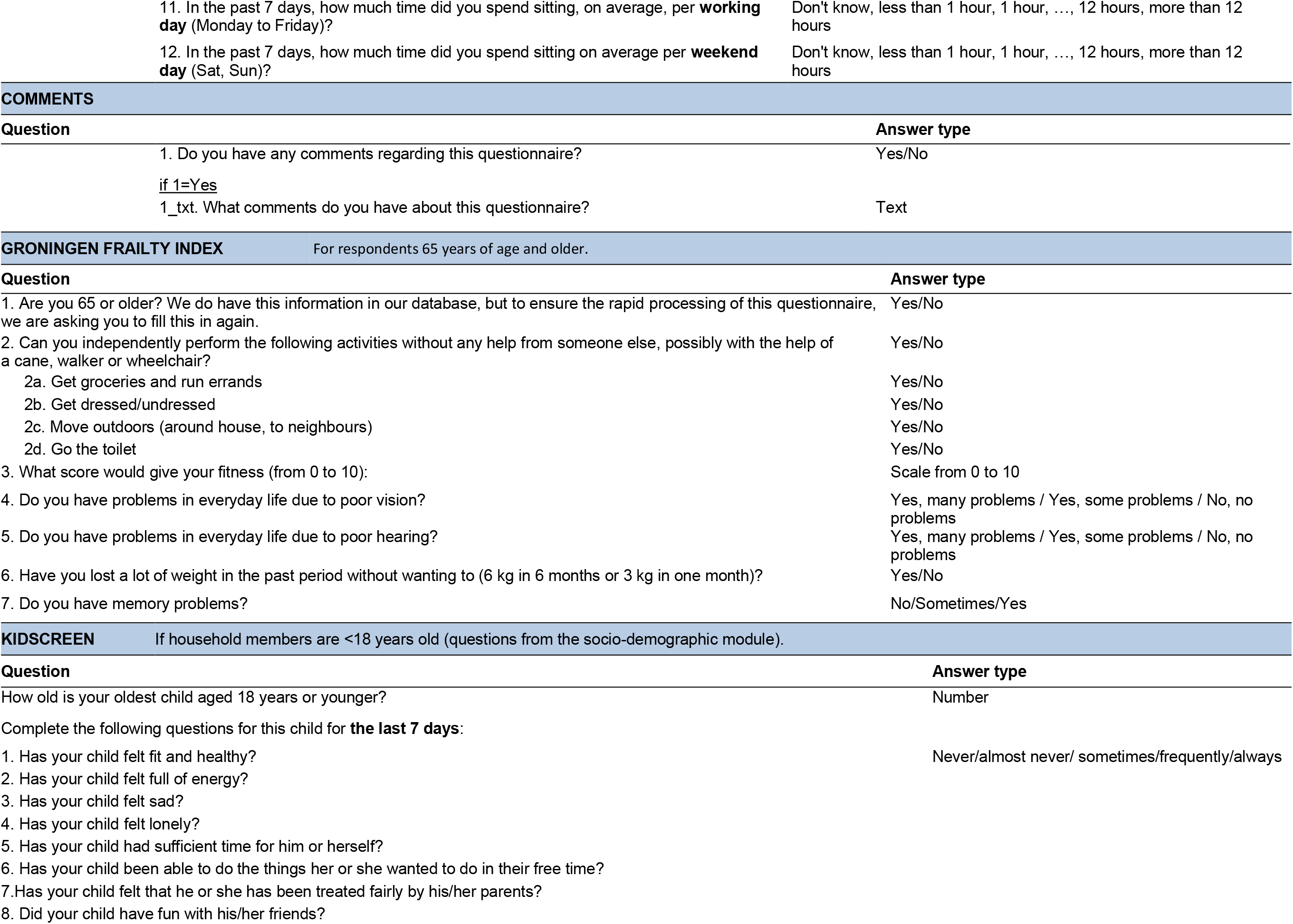
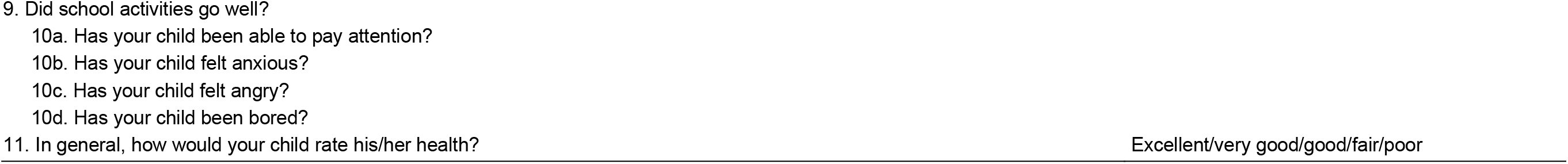
Lifelines COVID-19 questionnaire. Questions asked are modified slightly when respondents have filled in a previous questionnaire to indicate that they should answer with respect to the intervening period.

### Main questionnaire modules

#### Socio-demographics

Participants are asked about their household location, household make-up (number of co-habitants, number of children), work situation (employment status, employment location including working from home and whether they are still travelling to work, critical worker status and whether they are unemployed due to the crisis), the work situations of their household members, current weight, vaccination status (influenza and BMR) and pregnancy status. Participants who indicate they have children under 18 are asked additional questions about their children’s stress levels and experiences.

#### Chronic disease

Participants are asked whether they have a chronic health condition (cardiovascular disease, lung disease, kidney disease, diabetes, chronic muscle disease, psychiatric disorder, autoimmune disorder, cancer, neurological disease, spleen disease) and have the opportunity to elaborate if their condition is not on the specified list.

#### COVID-19

Participants are asked if they have tested positive for COVID-19 or been diagnosed with COVID-19 by a physician. They are also asked if they themselves think they have had COVID-19, if they know how they were infected, if any of their cohabitants have tested positive or been diagnosed, and if they have had physical contact with someone with COVID-19.

#### Health and symptoms

Participants are asked to assess their own health and to what extent they have experienced 28 specific symptoms in the preceding week (see Table 1 for full list). They are also asked to assess their experience of fatigue over the same time period.

#### Medication use

Participants are asked whether they have used any of eight classes of commonly used medications (see Table 1 for full list) and to indicate which medications. They can also input other medications not on the list.

#### Mental health and well-being

Participants are asked a series of questions to assess functioning of psychosomatic and mental health. To assess symptoms of depression and anxiety, a subset of the Mini International Neuropsychiatric Interview (MINI) [9] was included in the Lifelines COVID-19 questionnaire. The MINI has been administered in previous assessment waves of Lifelines to assess major depressive disorder (MD) and general anxiety disorder (GAD), which will allow for comparative analyses between pre- and post-outbreak. To evaluate a broader range of psychological problems and symptoms of psychopathology, the 12-item ordinal Symptom CheckList-90 Somatization subscale (SCL-90 SOM), which has been recommended for large-scale studies was surveyed [10]. The 12 items have five Likert-response options. To assess symptoms of fatigue, a subset of the checklist individual strength (CIS-20) [11] was included. Each CIS-20 item had seven Likert-response options. Both the SCL-90 SOM and CIS-20 were also previously implemented in Lifelines. Respondents further received questions on the impact of the pandemic on the mental health of their child or children, if applicable. These regarded three questions on stress of the child because of the Corona-crisis and feeling safe at home and in their community, for children aged 8-12 and 12-18 years.

#### Corona-related well-being

Participants are asked about their worries about the pandemic, what infection precautions they are taking, where they are getting their COVID-19 information, their perceptions of how the pandemic is affecting society and whether people close to them have had COVID-19 or died of COVID-19. They are also asked to rate their quality of life.

#### Social life & Connectedness

These two modules ask about feelings of isolation and exclusion and the extent to which participants feel connected to their families, neighbours, communities and nation.

#### Lifestyle

This module asks participants questions about changes in their eating patterns, exercise and activity levels and tobacco and alcohol use.

### Additional modules

#### COVID-19 and its impact on children

Parents participating in Lifelines NEXT also receive questionnaires related to COVID-19 and symptoms in children. While it is now thought that children rarely develop severe disease, and usually have the asymptomatic form, there are also reports of rare COVID-19 syndromes seen only in children [12]. Using these questionnaires, the project aims to look for the appearance of various symptoms in children. The questions are categorized into three age groups: questions for children aged 0-3 years, questions for children 4-7 years and questions for children 8-18 years old.

#### Supplemental question modules

Since the start of the Lifelines COVID-19 questionnaire programme, additional modules have been adopted. These include a module for participants 65 and older to examine the impact of the crisis on their daily functioning using the Groningen Frailty index [13] and a module for parents of children aged 8-18 that explores the impact of the crisis on the quality of life of children, measured by the KIDSCREEN-10 [14]. There is also now a questionnaire that examines the impact of COVID-19 on work participation and financial wellbeing, including measures on the quantity and quality of work performed, work absence, workplace and occupational characteristics. A module using the Positive and Negative Affects Schedule (PANAS) [15] was also recently added to the questionnaire.

## Preliminary results

### Response rates and characteristics of respondents

Based on the availability of an email address and being over 18 years of age, 139,713 out of 159,482 Lifelines participants are invited to respond to the COVID-19 questionnaire during each questionnaire round. Compared to non-invited subjects, these invited subjects were younger, slightly more often female, had a lower BMI and were more often never smokers (Table 2). Of the 139,713 Lifelines participants invited, 68,501 (49%) completed at least one of the questionnaires in the first 6 weeks of the programme. These responders were slightly older, slightly more often female, had a higher BMI and were less often current smokers and more often ex-smokers compared to non-responders (see Table 2). While Lifelines as whole has been shown to be representative of the regional population [7], these slight differences in cohort make up should be considered when looking at data from COVID-19 cohort. However, recruitment is still on-going, and the characteristics of the cohort may change over time.

**Table 2.** Characteristics of Lifelines participants invited to participate in the cohort and the participants in COVID-19 questionnaire cohort during the first six weeks of the project (Questionnaire rounds 1-6).

|  | **Invited** | **Not invited** | **P-value** | **Responded** | **Not Responded** | **P-value** |
| --- | --- | --- | --- | --- | --- | --- |
| N (%) | 139,713 (87.6) | 19,769 (12.4) |  | 68,501 (49.0) | 71,212 (51.0) |  |
| Current age, mean (sd) | 51.1 (13.6) | 57.8 (18.3) | < 0.01 | 54.3 (13.0) | 48.1 (13.5) | < 0.01 |
| Male sex, % | 41.6 | 42.3 | 0.07 | 39.2 | 44.0 | < 0.01 |
| BMI at last visit, mean (sd) | 25.9 (4.3) | 26.5 (4.9) | < 0.01 | 26.0 (4.3) | 25.8 (4.3) | < 0.01 |
| Smoking at last visit, % |  |  |  |  |  |  |
| never | 52.1 | 46.1 |  | 51.9 | 52.4 | < 0.01 |
| ex | 32.1 | 37.5 | < 0.01 | 34.1 | 28.8 |  |
| current | 15.8 | 16.5 |  | 14.0 | 18.8 |  |

**Table 3.** Characteristics of Lifelines NEXT participants invited to participate in the cohort and the participants in COVID-19 questionnaire cohort during the first six weeks of the project (Questionnaire rounds 1-6).

|  | **Invited** | **Responded** | **Not responded** |
| --- | --- | --- | --- |
| N (%) | 321 (100) | 159 (49.5) | 162 (50.5) |
| Current age, mean (sd) | 33.6 (4.9) | 33.0 (4.3) | 34.3 (5.3) |
| Male sex, % | 49.5 | 36.5 | 62.3 |

For Lifelines NEXT, 321 people were invited to participate in the first six weeks of the project, of whom 159 participated (49.5%). Compared to invitees, respondents were more likely to be female (73.5% of respondents versus 50.5% of invitees). As Lifelines NEXT recruits women who are currently pregnant, the age range was small and did not differ substantially between invitees, respondents and non-respondents. In Lifelines NEXT, 80% of all parents who responded to the main Lifelines COVID-19 questionnaire returned data on their children for the module on COVID-19 and its impact on children. In total, we have data for 76 children in week 5 of the COVID-19 questionnaire initiative: 64 children 0-3 years of age, 11 children 4-7 years of age and one child in the 8-18 age group.

### COVID-19 infections and the Menni et al. prediction model

Of the participants who responded in the first 6 weeks of the project, 1,034 (1.5%) responded that they had been tested for COVID-19, and 116 (0.2%) tested positive. In addition, 811 (1.2%) respondents said they had been told by a doctor that they probably had COVID-19, while 5,034 (7.3%) participants responded that they thought they had had COVID-19. Menni et al. have developed a model to predict positive COVID-19 cases based on symptoms reported [16], and we applied this model to the data from the questionnaire to predict COVID-19 cases within the cohort. Here we found that of all participants who were not tested for COVID-19, 6.1% were predicted to have had COVID-19. In participants who had been told by a doctor that they probably had COVID-19, 34.5% were predicted to have had COVID-19. In participants who responded that they thought they had had COVID-19, 22.0% were predicted to have had COVID-19. Of the remaining participants, 4.7% were predicted to have had COVID-19. The Menni et al model [16] had an AUC of 0.821 (95% CI: 0.765-0.878) when we applied it to our data using incident positive tests (n=49) and incident negative tests (n=408). The specificity was 0.828 and the negative predictive value was 0.944, indicating that the model is good at predicting the negative cases. However, the sensitivity and positive predictive value were much lower (0.592 and 0.293, respectively) and this indicates that for future applications, for example in genome-wide association studies, the ability of the model to accurately predict the true positive cases needs to be improved, and this work is on-going.

### Sharing of early results

Results of the study are continuously updated and shared with Lifelines COVID-19 cohort participants and the Dutch public through the Corona Barometer website (https://coronabarometer.nl/), which presents on-going results as interactive infographics (see snapshot in Figure 2), and through frequent social media posts and press releases. One of the earliest results of the project was a clear signal that feelings of loneliness and isolation were substantially stronger in individuals who lived alone and that this effect was strongest in the youngest age group of respondents (18-30 year olds, see corresponding panel in Figure 2). There was also an increase in the number of unemployed respondents who reported losing their jobs due the crisis, rising from 7.5% of unemployed respondents to the first questionnaire round (Q1) up to 14% by the week 5 questionnaire (Q5). More recent results have shown that by the end of May 2020, as the number of infections and hospitalizations dropped to low levels and schools and business reopened, respondents were reporting less anxiety, better sleep and fewer worries about losing their jobs.

## Current and future directions

The data collected from the Lifelines COVID-19 cohort is currently being analysed to address the four goals of the project. The COVID-19 cases reported by participants are being used to track the outbreak, and the symptoms reported by participants are being examined to generate a symptom-based COVID-19 prediction model, as described above. The data on chronic diseases, medication use and environmental factors (ranging from cohabitation to smoking) will be used to look for associations with SARS-CoV-2 susceptibility and COVID-19 severity, which can help identify risk factors, protective factors and comorbidities. While factors such as age, sex, BMI and certain chronic illnesses have now been associated with a more severe COVID-19 and higher mortality [17], there have also been questions about whether recent BMR vaccinations can be protective factors [18][19], and the data collected in this cohort should help address this question. Important questions about why pregnant women and children seem to be relatively protected will also be analysed. Finally, it will be possible to look at genetic factors in detail as 17,911 of the 68,501 COVID-19 participants who completed at least one questionnaire have been genotyped. Next steps include identifying participants who were also participants in the Lifelines cohorts for which we have more detailed data, e.g. participants with gut microbiome data, currently available for >10,000 Lifelines participants.

Mental health problems are known to increase in times of physical and psychological distress. The current COVID-19 pandemic is accompanied by strict government measures of social distancing and quarantine to contain and control the spread of the virus. As these events place significant stress on society and increase isolation and loneliness, close monitoring of mental well-being is important for both short- and long-term public health policies and individual-level care. Alertness in clinical systems and tailored mental health care may be needed during and after such a mass traumatic event. The data from the MINI MD and GAD modules and the societal impact modules of the questionnaire will allow researchers to i) longitudinally track the prevalence of symptoms and diagnoses of MD and GAD during the pandemic in the Lifelines and Lifelines NEXT populations, ii) associate symptoms with COVID-19 severity and outcome, iii) identify at-risk groups and individuals and iv) measure the impact of government policies on the overall mental health in the cohort.

The pandemic has had a major impact on the working lives of people in the Netherlands, and the questionnaire will help to address this impact. Healthcare workers are a particularly vulnerable group due to their higher risk of being infected by SARS-CoV-2, and for many workers in healthcare professions, the current working conditions include long work hours, cancelled holidays, working environments with adverse physical and psychosocial work conditions, i.e. high psychological and emotional demands and low control [20][21]. These working conditions, together with moral distress in relation to the family situation during lockdown, may increase the risk for mental health problems and sickness absence in this occupational group, which is dominated by women, and with a high baseline risk. Other “essential” occupational groups are also experiencing unprecedented changes in their working environments that may affect their physical and mental health as well as their labour market attachment. For many “non-essential” occupational groups that are now encouraged to work from home, the home working environment might not be suitable, as not all jobs can easily be done from home, and many families now have to combine working from home with caring for children. This will likely impact the productivity and quality of their work, as well as the level of stress.

The lockdown has led to a sudden disruption of the economy, and several economic sectors were effectively brought to a standstill. As a result, many workers in these sectors have been temporarily laid-off of work, while workers in other nonessential sectors were encouraged to work from home as much as possible. To protect people from losing their jobs and minimise the impact on self-employed people, the Dutch government implemented a series of financial measures. Despite these measures, however, a large number of workers who were already at a disadvantage because of flexible contracts lost their jobs shortly after the lockdown. The Lifelines COVID-19 questionnaire is monitoring changes in people’s current work situation by asking if they lost their job because of the crisis, if they are working in an essential job, and whether they have to work from home. The answers to these questions will be used to monitor both the impact of the crisis on the short- and longer-term labour market and to identify workers most at risk of losing their job. This is essential information for policymakers to be able to target measures to the most vulnerable groups in society and mitigate the financial impact of the crisis.

## Links to other national and international programs

All the data generated by the Lifelines COVID-19 questionnaire is linked to data held in Lifelines about its participants. This will allow for longer-term monitoring of participants beyond the timeline of the questionnaire program and the COVID-19 outbreak. Besides linkage to other Lifelines data, the Lifelines COVID-19 cohort data can also be linked to societal data held by Statistics Nederland, to drug prescription data held by IADB.nl via the Pharmlines initiative [22] and to SARS-CoV-2 testing data (including serological data) held by Certe and other Dutch laboratories. Overlap with enriched Lifelines datasets in Lifelines Deep [8], Lifelines DAG3 (an on-going project with metagenomic sequencing the gut microbiome of 10,000 Lifelines participants) and Lifelines NEXT [2] will also allow for research projects with a different scope. Given that there is now genetic data for 17,911 of the 68,501 COVID-19 cohort participants generated through the UMCG Genetics Lifelines Initiative (UGLI), it will be possible to look at genetic risk (and protective) factors. The Lifelines COVID-19 project is also participating in the COVID-19 Host Genetics Initiative [23], an international collaboration to share and analyse data to identify the genetic determinants of SARS-CoV-2 susceptibility, COVID-19 severity and outcomes.

The Lifelines COVID-19 questionnaire was designed to make comparisons with similar projects throughout Europe. Direct cross-national comparisons with projects in Denmark and France are possible, as they are using nearly identical questionnaires, and will provide unique opportunities to examine the effect of different governmental measures on mental health and well-being. This cohort is also part of COVID-MINDS, an initiative at UCL. These insights can be used to fine-tune some of the measures when there will be a second wave of the COVID-19 epidemic. Moreover, the Lifelines COVID-19 questionnaires have been requested by other (inter)national researchers as basis for designing their own questionnaires, i.e. separate research has been done on the experiences of COVID-19 patients, both hospitalised and not hospitalised, making use of the Lifelines COVID-19 questions.

## Strengths and limitations

One of the main strengths of this project is its embedding within the now long-running Lifelines prospective population cohort, which provides a rich data background about participants and the knowledge, infrastructure and relationship with participants necessary to recruit and engage participants during an evolving crisis. The high and sustained rate of response and the weekly questionnaires mean that the project will have a detailed longitudinal prospective view of both the outbreak and the wider psychological and societal impacts of the crisis. Another strength is the collaboration of researchers across a range of disciplines in designing and implementing the questionnaire, which means that the questions included can be used to address a wide range of research questions, can have immediate impact on policy and can be used to help design new policies to prevent and/or manage renewed outbreaks. Finally, Lifelines will continue to follow its participants for the coming decade and beyond, providing opportunities to examine the long-term health impacts of the pandemic.

The timing and nature of the COVID-19 outbreak in the Northern Netherlands, which diverged from that in other parts of the country, is both a strength and a limitation. The relatively low number of cases in the region, even accounting for undiagnosed cases, may seem to pose difficulties for statistical association analyses looking at COVID-19-related factors. However, even within our cohort, >800 participants have had a COVID-19 diagnosis, confirmed either through a positive test or through a doctor’s diagnosis, which permits a wide number of statistical association analyses. Moreover, the impact of the societal steps taken to reduce the rate of infection in more heavily impacted regions of the Netherlands and the impact of the associated economic crises should have similar psychological and social impacts in the Lifelines population. The fact that the outbreak in the North was effectively capped by public health steps now puts the questionnaire programme in an interesting position to monitor the immediate health and societal impacts of the lockdown measures and the subsequent impact of coming out of lockdown. It may also lay groundwork for steps to be taken if there is a later resurgence of COVID-19 infections, and the data generated while infection rates were low could work as baseline values if subsequent outbreaks in the Northern provinces are more intense.

## Collaboration

The data analysed in this study was obtained from the Lifelines biobank, under project application number ov20_0554. Researchers interested in using this data should contact the Lifelines Research Office.

## Data Availability

The data is available via the Lifelines biobank, a resource accessible for any researcher. For more information on the Lifelines biobank and data access, please visit https://www.lifelines.nl.

## Acknowledgements

We thank all Lifelines and Lifelines NEXT participants for repeatedly filling out our questionnaires and all the experts involved in developing the content of the questionnaires. We thank the Applied Health Research unit of the UMCG Department of Health Sciences, the UMCG Genomics Coordination Center, the UG Center for Information Technology and their sponsors BBMRI-NL & TarGet for storage and computing infrastructure. We thank Alex Friedrich and Gerolf de Boer for critical input for Figure 1 and supplying the R(t) data in the figure and Trishla Sinha for editing the Lifelines NEXT COVID-19 questionnaires.

---

**Supplementary Table 1.** COVID-19 infections, hospitalizations and deaths for the Netherlands as whole and for the Northern Provinces. Statistics are as of June 09, 2020. Source: RIVM, downloaded from the CoronaWatchNL Github.

| **Region** | **Infections** | **Hospitalizations** | **Deaths** |
| --- | --- | --- | --- |
| Netherlands | 47903 | 11800 | 6031 |
| North NL | 1491 | 320 | 122 |
| Groningen | 352 | 74 | 17 |
| Friesland | 616 | 130 | 65 |
| Drenthe | 523 | 116 | 40 |

